# The impact of trade and investment agreements on the implementation non-communicable disease policies, 2014-2019: protocol for a statistical study

**DOI:** 10.1101/2022.05.13.22274669

**Authors:** P. Barlow, L. Allen

## Abstract

**Introduction:** Regulating tobacco, alcohol, and unhealthy foods and drinks is a cornerstone of global efforts to combat the Non-Communicable Disease (NCD) pandemic, but implementation of these policies remains slow. It has been suggested that producers of these unhealthy commodities use rules in Trade and Investment Agreements (TIAs) to delay and undermine NCD policy implementation. Yet, there is no systematic empirical evidence linking TIA participation to reduced implementation. Here we present a study protocol for a statistical analysis of the relationship between TIA participation and the implementation of regulations on tobacco, alcohol, and unhealthy food and drink in 154 countries, 2014-2019.

**Methods and analysis:** We aim to examine whether participation in TIAs with the EU and US is associated with implementation of regulations targeting tobacco, alcohol, and unhealthy food and drink. We focus on TIAs with these countries as their TIAs create multiple opportunities to contest health regulations, and a majority of the major unhealthy commodity producers are registered in these jurisdictions. Partial and full implementation is captured in a recently published dataset which systematically coded implementation of 11 NCD policies in 2014, 2016 and 2019. We will combine these outcome data with TIA membership and covariate data from multiple sources. We will calculate descriptive statistics and use both regression adjustment and matching to conduct covariate-adjusted, quasi-experimental comparisons of implementation levels and progress according to whether or not countries have a TIA with the EU or US. Further analyses and robustness checks will examine additional TIA participation arrangements and test the sensitivity of our results to our model specifications.

**Ethics and dissemination:** Ethics approval will not be required as the study uses anonymised and pre-aggregated data. Findings will be disseminated to policymakers via personal contacts and press releases in parallel with scientific papers and conference presentations.

**Funding:** This research received no specific grant from any funding agency in the public, commercial or not-for-profit sectors.

## Background and study rationale

Non-Communicable Diseases (NCDs) including cardiovascular disease, chronic respiratory diseases, and diabetes kill 41 million people each year, equivalent to 71% of all deaths globally, and are considered a major challenge to sustainable development in the 21^st^ century (1,2). To tackle this global health crisis, WHO recommends governments implement a suite of ‘best-buy’ interventions, which are cost-effective means to prevent NCDs, including those summarised in Box 1 (3). Despite unanimous endorsement of these and other interventions targeting unhealthy commodities by all 194 WHO member states, implementation remains slow and uneven (4).

### Box 1.

**WHO-recommended ‘best-buy’ policies targeting the marketing, composition, and consumption of unhealthy commodities**

#### Tobacco

- Tobacco taxes
- Smoke-free place policies
- Graphic warnings on cigarette packages
- Tobacco advertising bans

#### Alcohol

- Alcohol sales or advertising restrictions
- Alcohol taxes

#### Unhealthy food and non-alcoholic beverages

- Legislation implementing the International Code of Marketing of Breast-milk Substitutes.
- Policies to reduce salt/sodium consumption
- Policies to limit saturated fatty acids and eliminate trans-fats
- Policies targeting the marketing of foods and non-alcoholic beverages to children

It has been suggested that the unhealthy commodity industries play a major role in delaying and undermining NCD policy implementation by invoking rules in Trade and Investment Agreements (TIAs) to contest regulations targeting the marketing and promotion of their products (5–9). TIAs include international investment agreements, which are treaties between two or more states for the purpose of promotion and protection of cross-border investments, and bi-lateral or regional free trade agreements, which seek to encourage both trade and investment (10). Between 1960 and 2020, governments worldwide ratified a total of 654 trade agreements and 2,841 investment treaties (11,12).

TIAs empower the unhealthy commodity industries to challenge, delay, and undermine regulations targeting tobacco, alcohol, and unhealthy food and drinks. This can occur as a result of specific commitments that governments make in TIAs, especially those within a new generation of ‘deep’ agreements (13). Deep TIAs expand investor protections and include ‘behind the border’ trade provisions which move beyond a historical focus on border taxes and quotas to harmonize or limit domestic regulations (11). One of the most controversial clauses establishes procedures for a form of international arbitration, referred to as investor-state dispute settlement (ISDS) (14). ISDS and other TIA commitments enable foreign corporations to strategically delay implementation of unhealthy commodity regulations by threatening or initiating a costly legal dispute about a policy (15,16). This delays implementation where governments back down in response to legal threats or disputes, a phenomenon known as ‘regulatory chill’ (15). In addition, deep TIAs can establish avenues for industry input in policy-making and commitments to sharing information about policy proposals (7). This creates opportunities for industry to contest regulations as they are being developed (17).

Delays in regulating unhealthy commodities are especially likely where governments sign TIAs with the US or EU as these countries regularly pursue deep TIAs, and many of the world’s largest and most profitable processed food, tobacco, and alcohol companies are headquartered in these jurisdictions (18,19). Deep TIAs in these jurisdictions bestow domestic companies with a potent means to stall implementation, whilst their substantial profits enable them to mount well-financed legal challenges that are prohibitively costly for many nations to withstand (20,21).

Researchers have identified several instances where multi-national businesses have invoked TIAs to oppose unhealthy commodity regulations. For example, the tobacco giant Philip Morris challenged the tobacco packaging legislation the governments of Australia and Uruguay had adopted by arguing the measures were inconsistent with the countries’ TIAs (8). These formal disputes received significant attention but were ultimately unsuccessful.

However, dispute threats and challenges made outside of courts occur more frequently and can be highly influential, as indicated in analyses of challenges to food, tobacco, soft-drink and alcohol regulations at WTO committee meetings (22). Case study research has identified several instances where these challenges and other dispute threats were followed by decisions to delay implementation (6,23).

Yet, the evidence linking industry pressure via TIA participation to actual policy outcomes remains weak. As far as we are aware, there is no quantitative evidence examining the relationship between TIA participation and policy implementation. Such an analysis requires systematic and harmonized data tracking policy implementation, or a lack thereof, across multiple years and countries. In the absence of such data, research has focussed on specific legal texts and descriptions of trade-related challenges or disputes without systematically connecting these to policy outcomes (17,22,24–27). Qualitative studies have also identified how concerns about legal threats and compliance with TIAs featured in policy-maker decision-making and suspected instances regulatory chill (28). Whilst previous research has been informative, it remains unclear whether TIAs have a causal impact on policy and whether select case studies are generalisable, or are instead high-profile exceptions to a broader tendency for governments to successfully implement policies despite their TIA arrangements and corresponding industry pressure.

These ambiguities are compounded by the recognition within most TIAs that governments have a legitimate need to regulate to protect public health (27,29). Such measures can be therefore deemed permissible according to the agreement, and governments that are aware of such flexibilities or who have a strong political commitment to NCD prevention can sometimes use these protections to withstand business challenges. For example, Chile’s successful defence of its front-of-pack labelling legislation against the food industry’s claim that the measure was incompatible with WTO rules has been partially attributed to the government’s access to legal expertise and its determination to withstand industry pressure (30). Furthermore, businesses operating in a jurisdiction with high levels of regulation may promote increased regulation elsewhere in order to avoid competitive disadvantages associated with higher domestic regulation (31). This possibility became evident when, in 2015, Nestle, Kellogs, Mars, Mondelez supported an EU-wide trans-fat limit in order to create “a common level-playing field for businesses” after similar legislation had been introduced in the US (32). TIAs may therefore promote policy convergence, with the net positive or negative effect dependent on the number and scope of NCD policies in TIA partner countries.

Here we outline a protocol for a statistical analysis of the relationship between TIA participation and the implementation of WHO-recommended regulations on unhealthy commodities. We use repeated cross-national data collected over 3 years (2014, 2016, and 2019) and statistical modelling to examine the relationship between TIA participation under different TIA arrangements, models, and assumptions.

## Objectives

The objective of this study is to examine whether participation in TIAs with the US or EU is associated with implementation of policies targeting tobacco, alcohol, and unhealthy foods and drinks. In sensitivity analyses we examine whether these associations apply specifically to US/EU TIAs, where TIAs are often ‘deep’ in scope and where many large unhealthy commodity producers are registered, or are instead generalizable to a range of other TIA participation arrangements.

## Methods

### Data and measurement

Our primary dependent variable of interest captures government implementation of policies targeting the marketing and promotion of tobacco, alcohol and unhealthy food and drink in 2014, 2016 and 2019. Implementation of these regulations is assessed by the WHO using regular NCD country capacity surveys. Cross-sectional implementation survey responses have been published for 194 countries reporting data from 2014, 2016, and 2019. We will use a recently published dataset which systematically coded WHO’s policy monitoring data across the three years as fully implemented, partially implemented, not implemented, or no data available. We subset this data to the 11 regulations targeting alcohol, tobacco, and unhealthy food and soft drinks, shown in Box 1.

For each of the 11 measures in Box 1, we create a dichotomous outcome variable indicating whether or not a county has achieved full implementation of the measure, coded as 0 (no implementation) or 1 (full implementation) in the NCD policy dataset. In subsequent analyses we examine whether our results are consistent when using alternative coding procedures (see ‘Additional Analyses’ below).

### Independent variable

Theoretically, implementation of the regulations outlined in Box 1 may be influenced by a range of TIA arrangements. In our primary analysis we examine the relationship between policy implementation and participation in US/EU TIAs, with secondary analyses examining alternative TIA participation indicators, as outlined below (see ‘Additional Analyses’). For our primary analysis we create a dichotomous ‘treatment’ variable indicating whether a country is a member of either a US or EU TIA (1) or not (0). We will create this variable using a list of investment treaties in force, by country-year, from the UN’s Investment Policy Hub (12). We complement this with data indicating trade agreements in force, by country-year, from the Design of Trade Agreements Database, DESTA (11).

### Statistical analyses

We will first use simple descriptive statistics to examine variation in implementation of unhealthy commodity regulations in countries with and without US/EU TIAs. We then estimate statistical models to adjust for potential confounding in these comparisons.

All statistical analyses will be performed in R. Statistical significance will assessed based on whether the p-value for each test is less than 0.05 (p<0.05). We will also report 95% confidence intervals.

### Descriptive analysis

Two-proportion z-tests will be used to assess differences in the proportion of countries that have achieved full implementation of each of the unhealthy commodity regulations in Box 1 according to whether or not they have TIAs with the US or EU. Z-tests will also be performed to compare the mean number of fully implemented regulations in each commodity category in countries with and without US/EU TIAs.

### Specifications to minimise confounding

Countries which do and do not participate in US/EU TIAs differ from one-another in ways that are associated with NCD policy implementation, for example with respect to democratization and GDP per capita (33,34). These differences may account for variation in policy implementation across countries with and without US/EU TIAs, as captured in the descriptive statistics above. To address this we adopt a quasi-experimental approach, which is appropriate for testing descriptive causal hypotheses when randomization is unfeasible, as is the case with TIAs (35).

There are multiple different ways to approach our analysis, and in order to obtain the most robust findings, we intend to employ three widely used statistical approaches that are feasible with our data, report diagnostic tests, and base on our main results on best performing models and most consistent results. The three approaches are:

1. regression adjustment,
2. matching, and
3. first-difference models.

Each approach has different advantages, disadvantages and data requirements (36–38). We will evaluate the performance of each model, and the main results presented in the paper will be those that are based on the best performing models with respect to statistical power and additional diagnostic tests, as outlined in further detail below. Data, code, and results from all models will be reported in supplementary appendices in order to mitigate against selective reporting.

#### Step 1: regression adjustment

Our baseline logistic regression model is:

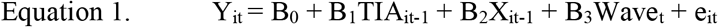

Where Y_it_ is the binary indicator of policy implementation of a given regulation in country i in year t (2014, 2016 or 2019), and B_0_ is the intercept. TIA_it_ is the indicator of US/EU TIA participation in country i in year t-1 with coefficient B_1_. To allow for a delayed effect, we lag this indicator by one year. X_it-1_ in Equation 1 is a vector of controls measured in year t-1 with coefficients in the vector B_2._ We control for variables known to be related to both treatment assignment and the outcome, and which are not affected by the treatment (39,40). We incorporate the following relevant and measurable covariates: democratization, GDP per capita, the share of the population of secondary education age that is enrolled in secondary education, implementation of non-trade business regulations, WTO participation, geographic region, participation in TIAs where other major unhealthy commodity producers are headquartered, and international political integration (or ‘political globalization’). Again these are lagged by 1 year in our models to allow for a delayed effect. Table 1 lists the data sources and measurement of these variables, and the rationale for their inclusion according to studies examining determinants of US/EU TIA ratification and NCD policy implementation (11,22,47,33,34,41–46).

**Table 1.** Covariates and measurement

| **Covariate** | **Rationale for inclusion** | **Measure, data type, range, and source** | **Data availability<sup>a</sup>** |
| --- | --- | --- | --- |
| Democratization | Democracies tend to have more liberal trade policies, and pairs of democracies are more likely to form TIAs. Democratization is also associated with higher NCD implementation scores.<br>Source: Mansfield and Milner (2012); Allen et al. (2020) | Varieties of Democracy (V-Dem) Multiplicative polyarchy index, which combines scores across multiple indicators of democracy (suffrage, free and fair elections, elected officials, freedom of civil and political organization, and freedom of expression).<br>Continuous (0-1), from V-Dem Institute, University of Gothenberg | 192-194 countries |
| GDP per capita | Countries with larger levels of GDP per capita tend to be more open, and countries with similar levels of GDP per capita and with are more likely to ratify TIAs. Countries with higher levels of GDP per capita also have more resources to implement NCD policies.<br>Sources: Egger, Egger & Greenaway (2008); Baier and Bergstrand (2009), Allen et al. (2020) | GDP per capita, PPP (constant 2017 international \$).<br>Continuous (731 to 155,201), from World Bank World Development Indicators | 178-194 countries |
| Human capital | Differences in skilled labour affect whether two countries ratify TIAs, whilst the school enrolment rate is a dimension of human capital that is associated with NCD policy implementation<br>Sources: Egger, Egger & Greenaway (2008); Allen (2020) | The ratio of total enrolment in secondary education (among persons of any age) to the population of the age group that is typically enrolled in secondary education.<br>Continuous (11.01 to 164), from World Bank World Development Indicators | 122-141 countries |
| Government stance on free market capitalism and liberalisation | Support for market liberalization and free markets is associated with trade liberalisation, whilst qualitative/ case-study research indicates that belief in the superiority of individual enterprise and preference against state intervention reduces political willingness to regulate unhealthy commodities | Orientation of the chief executive/ their party with respect to economic policy,<br>Nominal (Left, Right, or Centrist), from the Database of Political Institutions, 2020 edition https://mydata.iadb.org/ | 126-131 countries |
|  | Sources: Dutt and Mitra (2002), Milner and Judkins (2004), Thow et al. (2021), Barlow and Thow (2021) |  |  |
| Business regulations and domestic market liberalisation | Trade liberalisation is associated with other reforms to liberalise domestic markets, including limited business regulation, whilst support for minimal business regulation and domestic market liberalisation associates with opposition to, and limited implementation of, unhealthy commodity regulations<br>Sources: Billmeier and Nannicini (2013), Rodriguez and Rodrik (2001), Cullerton et al. (2016) | The extent to which the regulatory and infrastructure environments constrain the efficient operation of businesses (excluding trade policies), based on a weighted sum of 13 indicators in the World Bank's Doing Business report.<br>Continuous (0-100), from The Heritage Foundation | 186 countries |
| WTO Participation | WTO participation may incentivise RTA negotiation as governments seek to further liberalise trade (beyond what is possible within WTO), whilst WTO membership may independently influence NCD prevention<br>Sources: Baldwin 2009, Barlow et al. 2018 | WTO membership status Binary, from Head et al. (2020) and accessed via Centre d'Etudes Prospectives et d'Informations<br>Binary (0 or 1) | 194 countries |
| Participation in TIAs with other unhealthy commodity exporters (Switzerland, Australia, Canada, New Zealand) | Participation in US/EU TIAs associates with TIA ratification with other unhealthy commodity exporters (as, for example, the US/EU may want to secure the same market advantages as other states), whilst other unhealthy commodity exporters have been found to frequently challenge regulations targeting unhealthy foods, soft-drinks, tobacco and alcohol.<br>Sources: Barlow et al. 2018; Dur & Baccini 2014 | Binary indicator of participation in a TIA with Switzerland, Australia, Canada or New Zealand, as identified Bilateral Investment Treaties listed in the UN's Investment Policy Hub and FTAs in the Design of Trade Agreements Database, DESTA.<br>Binary (0 or 1) | 194 countries |
| International political integration | International political integration associates with exchange of information, norms and political pressure regarding both trade policy and NCD prevention, | KOF Political Globalisation index, calculated using the number of foreign embassies in a country, personnel contributed to UN security | 194 countries |
|  | potentially leading governments to seek and negotiate TIAs whilst also influencing policies targeting unhealthy commodities.<br>Sources: Dobbin et al. 2007; Valente et al. 2019 | council missions (%),the number of internationally oriented nongovernmental organisations (NGO) operating in that country, the number of multilateral treaties signed since 1945, the number of memberships in international organizations and a measure for the treaty partner diversity.<br>Continuous, 0-100.<br>From KOF Swiss Economic Institute |  |
| International economic integration | Countries with US/EU TIAs also tend to have larger trade and investment volumes (independent of the effects of US/EU TIAs) as, for example, the importance of trade to the national economy can lead countries to pursue TIAs, and countries may limit/ be pressured to limit policies targeting unhealthy commodities in an attempt to minimize trade disruptions.<br>Source: Baier & Bergstrand (2009); Barlow et al. 2018 | KOF de facto Economic Globalisation index, calculated using total trade in goods and services, trade diversity, FDI and portfolio investment, debt, reserves, and international income payments.<br>Continuous, 0-100.<br>From KOF Swiss Economic Institute. | 194 countries |
| Number of TIAs with other countries | Countries with US/ EU TIAs may also tend to have TIAs with other countries, reflecting a general support for trade and market-oriented policies, and these other TIAs may also contain clauses that businesses can make use of to pressure countries to change policies (so-called ‘forum shopping’), as their now global supply chains means they have may have operations in many countries.<br>Source: Busch (2007) | DESTA and UNCTAD (as per main variables)<br>Continuous | 194 countries |

Notes: a: all variables are lagged by one year, with the exception of secondary education for which we lag the variable by 2 years in 2020 due to limited data availability in 2019. Table shows data availability 2014, 2016 and 2019. Ranges show min to max in this period if not equal across years

Wave_t_ in Equation 1 is a control for the wave of data collection with coefficient B_2_. This adjusts for unobserved macro-economic and political factors which influence policy implementation, vary across data collection waves, and are common across all countries. e_it_ in Equation 1 is the error term. We estimate Equation 1 using pooled logistic regression models and account for within-country correlations in the error term by clustering the standard errors at the country level.

Finally, we use the estimated coefficients in this model to calculate predicted probabilities of full implementation of each regulation in countries with and without US/EU TIAs as well as the Average Marginal Effect of US/EU TIAs, that is, the difference in theses predicted probabilities (48).

#### Step 2: matching

Regression adjustment does not fully correct for imbalance in covariates across countries with different TIA arrangements and can lead to inferences that are beyond the bounds of what is observed within the data (49,50). Matching has been widely applied to address these issues in analyses of the economic, environmental, and health consequences of TIAs (33,51–54). Matching pre-processes the data to identify the ‘untreated’ comparison country or countries that is/are most similar to each country with a EU/US TIA, thereby reducing imbalance in covariates (40). After the matching, the quality of matches and post-maching covariate balances can be analysed using a range of diagnostic tests. The matched sample is then analysed using regression models with controls for any remaining covariate imbalances (55).

There are many different matching algorithms, and we plan to employ the multiple procedures that are feasible with our data, report diagnostic tests, and base on our main results on best performing models and most consistent results (40). When first assessing ‘similarity’ between countries to identify relevant matches, we begin with one of the most commonly used, unit-free metrics: the propensity score. We estimate a logit model to predict treatment assignment (ie US/EU TIA participation) based on the set of covariates used to estimate Equation 1. Predicted probabilities of US/EU TIA participation are then calculated from this model; these are the ‘propensity scores’. We then use three alternatives to traditional propensity score matching: i) the non-parametric covariate-balancing propensity score algorithm, which identifies propensity scores such that both covariate balance and prediction of treatment assignment are maximized, and ii) calculation of an alternative measure of similarity/dissimilarity, the Mahalonobis distance.

Next, for each procedure, we select comparison countries based on whether their propensity scores are similar to the ‘treated’ countries with a US/EU TIAs. We use ‘nearest-neighbour’ matching to identify the 1- and 3-, and 5-most similar untreated countries. Since nearest matches on propensity scores may nevertheless be dissimilar to the countries with US/EU TIAs, we will set a caliper defining the maximum difference in propensity score to make a country match (‘radius matching’); in our case, we will use a caliper of 0.1 (i.e., ten percentage points of the likelihood of becoming a US/EU TIA member). This choice is necessarily arbitrary, and we also use alternative caliper specifications and choose these where they yield better performance on our diagnostic tests (see below).

As stated above, diagnostic tests will be used to determine which of the above models performs best and hence is to be used to estimate US/EU TIA impacts on policy. Following recommendations in Stuart (2010) and King and Zeng (2006), these tests include examination of the number of successfully matched pairs, covariate balance tests, comparing the difference in covariate means and variance, and tests of common support which examine the distribution of propensity scores across treated/untreated units and the ‘convex hull’ of the covariates (40,50). We will include all diagnostic test results in the study write-up and document the rationale for our final selection.

Finally, as a robustness check we will weight observations in inverse proportion to their similarity in the propensity score (as calculated from the best performing model above), rather than select a limited number of comparison countries, so that more ‘untreated’ units are used but more dissimilar units are given lower weight (‘kernel matching’). This approach increases statistical power, which can be limited with the aforementioned approaches, although it comes at a cost of incorporating some comparison countries which may be dissimilar to the countries with EU/US TIAs.

#### Step 3: first difference models

The models described above control for measurable confounding/covariate imbalance but do not address the influence of unobserved or unmeasured factors that predict both US/EU TIA participation and NCD policy implementation. A common strategy to reduce the influence of time-invariant unobserved/unmeasurable confounders is to estimate fixed-effects models (38). However, with only 3 years of data we lack sufficient data to estimate sufficiently powered fixed-effects models. We nevertheless exploit the temporal nature of our data by modelling changes over time in the outcome between two data collection waves (2015-2017 and 2017-2020). According to An and Winship (2015), this strategy is advantageous when few years of data are available as first differencing the outcomes helps to remove the influence of unobserved time-invariant factors, whilst posing fewer data restrictions than fixed-effects models. This approach is therefore recommended by An and Winship and has previously been applied to study political and macro-economic influences on health with few years of data (56–58).

After first-differencing the outcome, we apply the matching and weighting procedures outlined above on the differenced outcomes, modelling changes in implementation between two periods as a function of US/EU TIA participation and covariates in the first period.

### Patient and Public Involvement

No patient involved.

## Additional analyses

We conduct additional analyses to examine heterogeneity in the relationship between TIA participation and unhealthy commodity regulations across countries. We further conduct a number of tests which evaluate i) whether our results are consistent using alternative outcome and TIA participation indicators, ii) the specificity of our results, and iii) whether our results are robust when using alternative model specifications.

### Heterogeneity analyses

Our heterogeneity analyses examine whether the relationships examined above vary depending on the income level of the country and state capacity (indicators in Table 2 below), both of which have previously been identified as important modifiers of the relationship between TIAs and domestic health policy (20,59). We also examine if our results pertain specifically to US TIAs, since US companies have regularly been cited as the main actors that used strategic litigation to oppose NCD policies (8,26,60). For comparison we will conduct a separate analysis to assess if our results pertain specifically to EU TIAs.

**Table 2.** Data sources for additional analyses

| **Covariate** | **Measure, data type, and data source** | **Data availability** |
| --- | --- | --- |
| Country-income group | Income classification according to World Bank (based on GNI/capita thresholds)<br>Ordinal (low, lower-middle, upper-middle, and high-income)<br>From World Bank World Development Indicators | 194 countries |
| State capacity | Composite indicator of the perceptions of the quality of public services, civil service, policy formulation, implementation, and the credibility of the government's commitment to such policies.<br>Continuous indicator, measured as a percentile rank indicating the country's rank among all countries covered by the aggregate indicator, with 0 corresponding to lowest rank, and 100 to highest rank.<br>Originally developed by Kaufman et al. (2011), obtained via World Bank API | 194 countries |
| Distance between potential TIA partners | Great circle distance between country economic centers<br>Continuous, in km (1 – 19,951)<br>From Head et al. (2020), accessed via Centre d'Etudes Prospectives et d'Informations | 194 countries |
| Common border between potential TIA partners | Binary indicator of sharing a common border<br>Binary<br>From Head et al. (2020), accessed via Centre d'Etudes Prospectives et d'Informations | 194 countries |
| Whether potential TIA partners share a common language | Binary indicator of sharing a common language<br>Binary<br>From Head et al. (2020), accessed via Centre d'Etudes Prospectives et d'Informations | 194 countries |
| Implementation of NCD policies not targeting unhealthy commodities | Number of NCD policies not targeting unhealthy policies that achieved partial/ full implementation<br>Continuous<br>From Allen et al. (2020, 2021) | 194 countries |

Notes: a: all variables are lagged by one year, with the exception of international political integration, for which we lag the variable by 2 years in 2020 as no data are available in

### Alternative TIA participation measures

As noted above, implementation of unhealthy commodity regulations may be influenced by a range of TIA arrangements and the relationships we examine above may not apply exclusively to US/EU TIAs. We therefore re-estimate the relationship between TIA participation and unhealthy commodity regulations using several alternative TIA participation indicators. As with our main models we use regression adjustment, matching, and first-difference models to perform these tests. However, some additional analyses examine continuous rather than binary indicators of TIA participation (e.g. number of TIAs rather than participation in either a US or EU TIA). Since matching is not appropriate for analyses of continuous treatment indicators, we use an alternative but similar approach to address covariate imbalance: non-parametric Covariate Balancing Generalized Propensity Score (npCBGPS) estimation (61). npCBGPS uses an algorithm to search for a set of country-weights which, when applied to the data, minimises the correlation between the covariates and the probability of a given number of TIAs, whilst simultaneously maximising treatment prediction. We then apply these weights to the data when estimating our regression models. We estimate ‘doubly robust’ regression models, which incorporate the weights as well as the regression controls specified above (62).

We first re-estimate our models replacing the indicator of participation in a US/EU TIA with the following:

i. The *total number* of TIAs with the US and EU that a country participates in,
ii. The number of TIAs *with the US only* that a country participates in,
iii. The number of TIAs *with the EU only* that a country participates in.

Second, we examine whether our results apply to all TIAs with countries that are major producers unhealthy commodities, rather than the US/EU specifically. To assess this we will create variables which capture whether or not a country has a TIA with at least one other country where a major producer of unhealthy commodities is registered, with a second variable indicating the number of these TIAs. To identify these countries, we will use data from the 2014, 2016 and 2019 *Forbes* Global 2000 reports showing where the 10 largest, publicly listed companies in the alcohol, tobacco, and food/soft-drink sectors were registered in those years (63). We will create three separate ‘treatment’ (TIA participation) variables when examining implementation of policies targeting each category of commodities: i) tobacco, ii) alcohol, and iii) unhealthy food & drink. For each commodity, we code TIA participation as ‘1’ if a country has at least one TIA with a country in which one of these largest top 10 producers of the commodity in question is headquartered, or 0 otherwise. In additional analyses we will count the number of TIAs with these countries to assess how this is associated with unhealthy commodity regulation.

Third, we examine the total number of TIAs a country participates in, which creates exposure to multi-national companies operating in multiple jurisdictions. In addition, having multiple TIAs creates exposure to ‘venue’ or ‘forum shopping’, whereupon businesses use the TIA which is most advantageous to their case (6,64,65). We assess this by calculating the total number of TIAs in each country in each year (2014, 2016 and 2019). In subsequent analyses we interact the number of TIAs with the number of TIA partners to examine variation according to the number of distinct TIA partners, as several countries have multiple agreements with one partner country.

Finally, there is likely to be a strong correlation between each of the TIA participation indicators described above (for example participation in US TIAs is associated with the total number of TIAs), and so our analysis of one indicator may capture the influence of another. Furthermore, the association between each indicator and policy implementation may vary in magnitude. We therefore estimate a final model in which we incorporate the following TIA participation indicators simultaneously in order to adjust for this co-variation and compare the magnitude of their estimated impacts: total number of TIAs, US TIA participation, EU TIA participation, and participation in a TIA with a major producer of the unhealthy commodity in question. This model also adjusts for the same covariates as our baseline model, as described above.

### Alternative outcome measures

We examine several alternative outcome indicators. As noted above, TIAs could promote policy convergence, leading to more similar regulations among TIA partners, and whether this leads to a lower or higher level of regulation may depend on the specific regulations in force in TIA partner countries. In a first set of analyses we re-estimate our models substituting the outcome variable with an indicator of similarity in implementation in a given country compared to the mean in the TIA partner countries (the EU, US, or across all TIA partners, depending on the TIA participation indicator being evaluated). This indicator of similarity in implementation is calculated by dividing the implementation score in TIA partner countries with the implementation score in a given country with (or without) a TIA, so that values closer to 1 indicate more similar implementation levels.

We further re-estimate the above models using an alternative indicator capturing either no implementation (0), or at least partial implementation, i.e., coded as partial or full implementation in the NCD policy dataset (1), rather than full implementation only. We will further count the number of partially/fully implemented regulations across all commodities and each category of commodities, and examine the association between participation in US/EU TIAs and the number of regulations partially/fully implemented using a Poisson regression model.

Finally, we will create variables capturing whether countries move from no to partial/full implementation, or from partial to full implementation, between 2014 and 2016 and 2016 and 2019, and examine the association between participation in US/EU TIAs and each of these variables.

### Sensitivity analyses

We first conduct analyses to assess whether our results are sensitive to our propensity score model specification and our decision to use this metric rather than alternative measures of similarity between countries (40,66). First, Stuart (2010) recommends choosing a parsimonious list of variables when estimating propensity score models in small samples like ours, as including a large number of variables can lead to increases in variance. However, the results from our matching models may be sensitive to our choice of covariates and limited selection thereof. We therefore conduct a test in which we incorporate additional variables that predict US or EU TIA participation: distance, sharing a common border, sharing a common language. We also re-estimate all models (regression and matching) incorporating a further possible covariates of TIA formation and NCD policy to assess the sensitivity of all models to the inclusion of these additional variables: the total number of TIAs in force in a country, and social globalization (67).

Second, matching on the propensity score has the advantage of using a single indicator to perform matches, rather than multiple variables across which matches may not be possible, a phenomenon known as the ‘curse of dimensionality’. However, a disadvantage of propensity score matching is that covariate imbalances may not be sufficiently reduced (40,66). We therefore conduct an additional analysis in which we use an alternative metric recommended by King and Nielsen: the Mahalonobis distance. This metric is calculated by taking a weighted-measure of similarity in covariates in treated and untreated units.

Next, we assess whether our results may be attributable to alternative explanations. We re-estimate our models with a control for the number of fully implemented measures in the NCD policy dataset that did not directly regulate unhealthy commodities. This control captures other influences on implementation which may bias our estimates. We further conduct a placebo analysis, in which we re-estimate our models examining implementation of an NCD policy that we would not expect to be affected by TIAs: whether or not a country has set time-bound national targets to address NCDs based on WHO guidance. This helps to identify whether our results are driven by TIA implementation or some third factor leading to general changes in implementation.

Table 2 below lists the data sources and measurement of the variables included in our additional analyses.

### Missing data

For all analyses outlined above we will apply listwise deletion where data are missing. However, this approach can produce biased results where data are missing not at random (68). In addition, listwise deletion markedly reduces the sample size, which in turn causes a substantial loss of precision and power. Finally, the developers of the NCD policy implementation dataset assigned a score of zero for those implementation policies that are not reported on the basis that if a country is unable to determine whether a policy has been implemented, then it is likely that the policy was not implemented. However, it remains possible that assigning a score of zero in such cases will exert a downward bias on our estimates (for example if countries successfully implemented policies and were coded as zero in the data).

To address these limitations we will re-estimate our models using imputed outcome and covariate data. To impute these missing data we will use a multivariate normal model (with log transformations for non-normal variables) incorporating all covariates in our original model. The number of imputations will follow von Hippel’s rule of thumb: that the number of imputations (m) should be similar to the percentage of cases that are incomplete. Hence m will be determined based on the number of missing cases in the data (69). We will then re-estimate our models using the imputed datasets and combine the results from these models, using Rubin’s Rules to calculate the Standard Errors.

## Ethics and dissemination

Ethical approval is not necessary as all data are publicly available. Furthermore, all data are collected at the country-level and it is not possible to identify individuals in these aggregated data. Findings will be disseminated via scientific papers and conference presentations.

## Conclusion

Using a novel dataset capturing NCD policy implementation and a range of statistical models, this study will provide new insights into the relationship between TIA participation and implementation of regulations targeting unhealthy commodities. The findings will help researchers and policymakers better understand whether TIAs actually constrain implementation of regulations targeting unhealthy commodities, and hence whether TIAs may need to be modified in order to facilitate government efforts to curb the global non-communicable disease pandemic.

## Data Availability

All data produced will be available upon reasonable request to the authors

